# Differential Redistribution of Activated Monocyte and Dendritic Cell Subsets to the Lung Associates with Severity of COVID-19

**DOI:** 10.1101/2020.05.13.20100925

**Authors:** Ildefonso Sánchez-Cerrillo, Pedro Landete, Beatriz Aldave, Santiago Sánchez-Alonso, Ana Sánchez Azofra, Ana Marcos-Jiménez, Elena Ávalos, Ana Alcaraz-Serna, Ignacio de los Santos, Tamara Mateu-Albero, Laura Esparcia, Celia López-Sanz, Pedro Martínez-Fleta, Ligia Gabrie, Luciana del Campo Guerola, María José Calzada, Isidoro González-Álvaro, Arantzazu Alfranca, Francisco Sánchez-Madrid, Cecilia Muñoz-Calleja, Joan B Soriano, Julio Ancochea, Enrique Martín-Gayo, on behalf of REINMUN-COVID and EDEPIMIC groups

## Abstract

The SARS-CoV-2 is responsible for the pandemic COVID-19 in infected individuals, who can either exhibit mild symptoms or progress towards a life-threatening acute respiratory distress syndrome (ARDS). It is known that exacerbated inflammation and dysregulated immune responses involving T and myeloid cells occur in COVID-19 patients with severe clinical progression. However, the differential contribution of specific subsets of dendritic cells and monocytes to ARDS is still poorly understood. In addition, the role of CD8^+^ T cells present in the lung of COVID-19 patients and relevant for viral control has not been characterized. With the aim to improve the knowledge in this area, we developed a cross-sectional study, in which we have studied the frequencies and activation profiles of dendritic cells and monocytes present in the blood of COVID-19 patients with different clinical severity in comparison with healthy control individuals. Furthermore, these subpopulations and their association with antiviral effector CD8^+^ T cell subsets were also characterized in lung infiltrates from critical COVID-19 patients. Collectively, our results suggest that inflammatory transitional and non-classical monocytes preferentially migrate from blood to lungs in patients with severe COVID-19. CD1c^+^ conventional dendritic cells also followed this pattern, whereas CD141^+^ conventional and CD123^hi^ plasmacytoid dendritic cells were depleted from blood but were absent in the lungs. Thus, this study increases the knowledge on the pathogenesis of COVID-19 disease and could be useful for the design of therapeutic strategies to fight SARS-CoV-2 infection.

**Single-sentence summary:** Depletion from the blood and differential activation patterns of inflammatory monocytes and CD1c^+^ conventional dendritic cells associate with development of ARDS in COVID-19 patients.

## Introduction

The new SARS-CoV-2 virus causing the COVID-19 (coronavirus disease 2019), has unleashed the current pandemic. Individuals infected with this pathogen can either remain asymptomatic or progress from mild to severe clinical conditions that include the acute respiratory distress syndrome (ARDS) and death. Analytical parameters such as high concentrations of IL-6 and acute phase reactants in plasma correlate with the development of severe symptoms in COVID-19 patients (1, 2), suggesting dysregulated immune responses in these individuals. Indeed, a cytokine storm-related syndrome has been proposed as a trigger of ARDS (3, 4) and in accordance, treatments to control inflammatory cytokine signaling are being used to reduce mortality of COVID-19 patients (5, 6). However, it is unknown whether specific subsets of innate and adaptive immune cells could be differentially contributing to a dysregulated immune response underlying the development of ARDS in COVID-19. The most recent studies indicate that immune exhaustion of effector T lymphocytes and altered humoral responses (7–9), combined with alterations in myeloid cells like monocytes (10, 11) might be related to increased inflammation and contribute to disease progression in SARS-CoV-2 infected patients. Dendritic cells (DC) are a heterogeneous lineage of antigen presenting cells (APC), which includes different subsets of CD123^hi^ plasmacytoid DCs (pDC) and conventional (cDC) CD1c^+^ and CD141^+^ DCs. These innate cells are critical for activation of adaptive CD4^+^ and CD8^+^ T cell responses, and are key players in the immune responses to viral and bacterial infections (12–16). A second important player in the immune response against pathogens are monocytes (Mo), which can be subdivided in immature classical (CD14^++^CD16^−^, C) and more differentiated inflammatory transitional (CD14^+^CD16^+^, T) and non-classical (CD14^−^CD16^++^, NC) subsets. Altered homeostasis of these specific populations has been linked to chronic inflammation and autoimmunity (17–19). Therefore, it is critical to assess in depth myeloid cell populations in COVID-19 to generate new knowledge that may contribute to the development of effective specific treatments. Additionally, it is important to better understand adaptive antiviral responses potentially present in COVID-19 patients, and how they might be associated with altered myeloid cells. Recent studies suggest that exhausted CD4^+^ T cell responses might be dysregulated in COVID-19 patients (20), but less is known about antiviral CD8^+^ T cells, which could be important for virus control. Assessment of a small number of COVID-19 patients suggests that CD8^+^ T cells infiltrate lungs during the course of the disease, but little information is available about which specific effector subsets are recruited (9). Therefore, an integrative view of the myeloid and lymphoid populations altered in peripheral blood and lung tissue of the patients and their association to the severity of COVID-19, is clearly needed. In the present study, we contribute to this aim by analyzing the frequencies and activation profiles of different DC and Mo subsets present in the blood of COVID-19 patients with different levels of clinical severity and by identifying cell subsets associated with protection or disease progression. Moreover, we have characterized inflammatory Mo and DC subsets as well as effector CD8^+^ T lymphocytes that are specifically recruited to the lung during progressive ARDS.

## Results

### Inflammatory patterns and severity define subgroups of COVID-19 patients

Our aim is to evaluate the impact of SARS-CoV-2 infection in circulating myeloid populations. To this end, we followed a cross-sectional study strategy (See STROBE flow chart, Fig. S1) and collected peripheral blood samples from a total of n = 64 individuals with COVID-19 (Median age 61 (min-max, 22–89), 57% male; 95% receiving treatment) who were recruited after a median of 3 days (0–25) upon admission (Table S1). For comparison purposes, 22 healthy donors were included in the study (Table S1). To correlate particular profiles of frequencies and activation of peripheral blood inflammatory cells and the severity of the disease, we stratified our cohort of COVID-19 patients into 3 groups of mild (G1), severe (G2) and critical (G3) disease, following recently described criteria (21). Their demographic and clinical characteristics are summarized in Table S1 and Table S2. As shown in Fig. S1 and in agreement with previous studies (2), we observed a significant and progressive decrease of PaO2/FiO2 (PaFiO2) (p<0.0001 for G1 vs G3) and increase of plasma IL-6 (p<0.0001 for G1 vs G3) and other inflammatory parameters such as Procalcitonin (PCT, p = 0.0008 G1 vs G3) and C Reactive Protein (CRP, p<0.0001 G1 vs G3) that was correlated with COVID-19 severity (Fig. S1).

### Differential redistribution of myeloid cell subsets in the blood of COVID-19 patients is associated with severity

To evaluate the impact of SARS-CoV-2 infection in circulating myeloid populations, we next examined whether specific myeloid subsets could be differentially altered in the three groups of COVID-19 patients and associated with disease progression. Using a multi-color flow cytometry panel, we determined the frequencies of the following circulating myeloid cell subsets: C Mo, T Mo, NC Mo, granulocytes, CD1c^+^ and CD141^+^cDC and CD123^hi^ pDC (for gating strategy, see Fig. S2). Using a tSNE tool, we were able to observe differences in mononuclear myeloid cells distribution based on the abundance of cell populations between COVID-19 and healthy control individuals (Fig.1A left). A more detailed analysis suggested that these changes were due to alterations on specific myeloid subsets in COVID-19 individuals (Fig.1A, right). An individual analysis of each myeloid subset in blood showed a significant reduction of almost all circulating myeloid cell subsets when considering all COVID-19 patients (Fig. 1B). Interestingly, certain myeloid populations were more significantly affected in the different patient subgroups. In particular, CD123^hi^ pDC and CD141^+^ cDCs were significantly diminished in the three patient groups, suggesting a disease-rather than severity-related impact on these cells (Fig. 1B). On the other hand, the decrease of CD1c^+^ cDCs, and C and NC Mo was more pronounced in severe G2 and critical G3 COVID-19 patients and therefore indicating an association with severity (Fig. 1B). In contrast, T Mo followed a completely different pattern, since they were significantly increased in mild G1 COVID-19 patients compared to healthy controls but dramatically decreased in critical G3 COVID-19 patients (Fig.1B). Moreover, T Mo/CD1c^+^ cDCs ratios were higher in G1 and G2 patients with less severe disease progression (Fig. S2C). In addition, big sized CD14^−^ CD16^hi^ HLADR^−^ granulocytes were increased in G1 and G2, but not G3 COVID-19 subgroups; and higher ratios of these cells relative to CD1c^+^ DCs were specifically increased in severe G2 patients (Fig.S2B-2C). Alterations in frequencies of specific myeloid populations have been related to clinical parameters associated with COVID-19 severity (2). Our analyses revealed that higher frequencies of circulating C, T and NC Mo (Fig. 1C) and CD141^+^ cDCs (Fig. S3B) were moderately correlated with higher levels of blood oxygenation. Importantly, depletion of C and T Mo from the blood correlated most significantly with higher plasma levels of inflammatory markers such as PCT, CRP and in the case of C and T Mo, with higher IL-6 (Fig. 1C-D, Fig.S3A). Similar trends were observed for cDC (Fig. S3A), while no significant association was observed for granulocytes (Fig. S3B). On the other hand, depletion of inflammatory NC Mo in COVID-19 did not associate with IL-6 plasma levels (Fig.S3A). Together, these data indicate that frequencies of C, T and NC Mo and CD1c^+^ cDCs are preferentially reduced in the blood during severe disease progression and these populations are differentially correlated with inflammatory markers.

**Figure 1.**
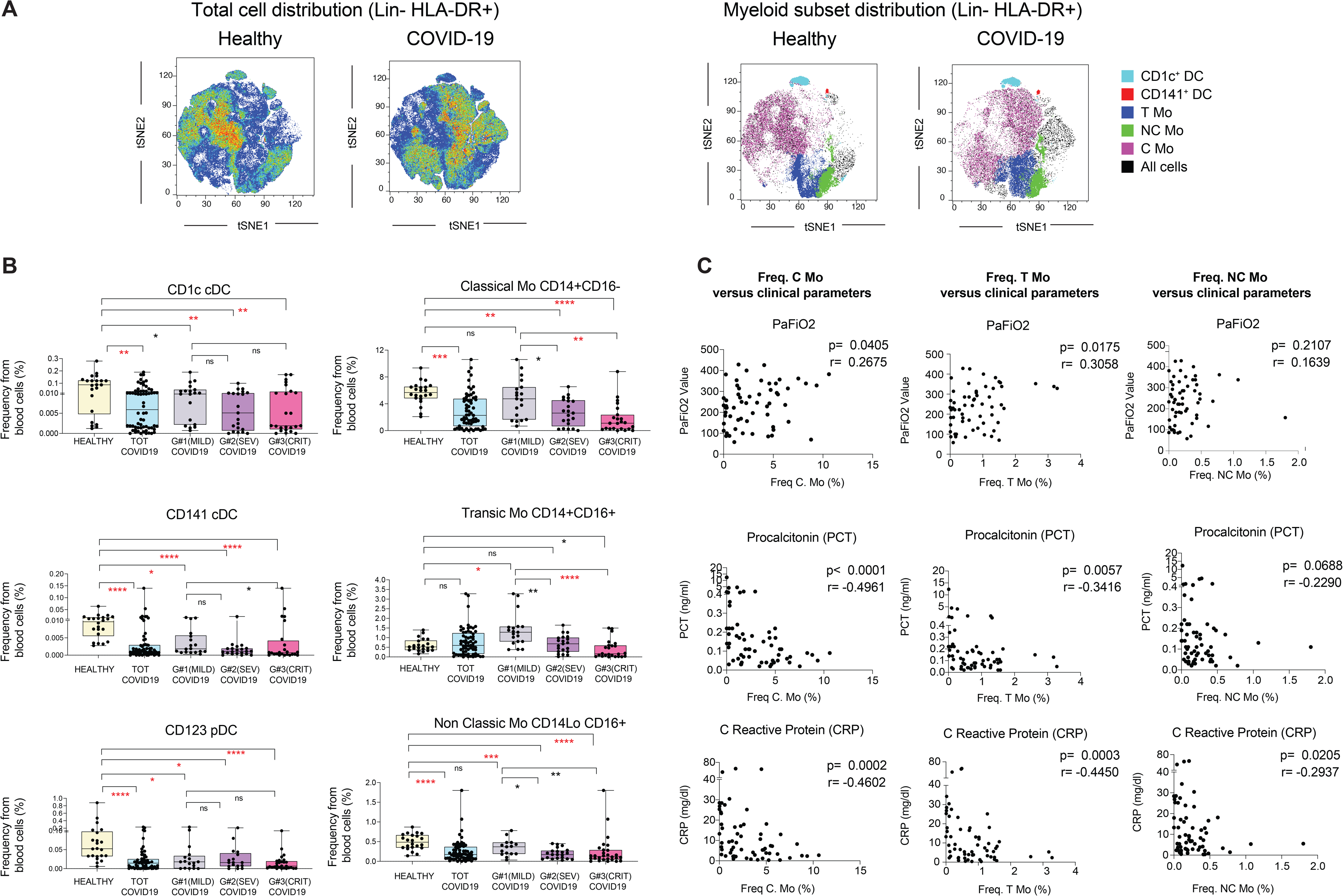
Analysis of different myeloid subsets in the blood of COVID-19 patients with different clinical severity. (A): tSNE analysis of myeloid cells from a total of 49 samples (34 COVID-19 patients and 15 healthy controls) gated after exclusion of lineage positive cells and excluding granulocytes. Plots on the left show combined density of cell clusters in both patient groups. Plots on the right display highlighted distribution of each indicated myeloid cell population. (B): Proportions of indicated myeloid cell populations present in the blood of healthy individuals versus either total COVID-19 patients included in the study or patients stratified into groups according to mild (G1), severe (SEV, G2) and critical (CRIT, G3) clinical status as shown in Table S1. Statistical significance of differences between patient groups was calculated using a nonparametric two tailed Mann Whitney test (black). Comparisons that remained significant after a Kruskal Wallis test followed by a Dunn’s post hoc-test for multiple comparisons are highlighted in red. *p<0.05; **p<0.01; ***p<0.001; ****p<0.0001. (C): Spearman correlations between frequencies of transitional (left) and non-classical (right) Mo and values of PaFiO2 (top plots), Procalcitonin (PCT, middle plots) and C reactive protein (CRP) detected in the blood of all COVID-19 patients included in the study. P and R values are shown in the upper right corner on each plot.

### Activation profiles in circulating monocytes versus dendritic cells from mild, severe and critical COVID-19 patients

We next sought to determine whether the activation state of DCs and Mo subsets was different in the subgroups of COVID-19 patients. Our previous tSNE analysis involved redistribution of cell populations considering both frequencies and expression of the maturation marker CD40, which has been linked to cellular activation and the expression of IL-6 cytokine (22, 23). As shown in Fig.2A, tSNE distribution of CD40 expression was restricted to specific myeloid cell populations while other cells seemed to have downregulated this molecule on COVID-19 patients (Fig. 2A). When CD40 expression levels were analyzed within each myeloid cell subset in total and stratified COVID-19 patients, we observed again differences associated with disease severity in some populations (Fig. 2B). Interestingly, CD40 expression tended to be lower in all Mo subsets from G3 patients, but most significantly on T and NC Mo (Fig. 2B). In contrast, no significant changes in CD40 expression were observed for CD1c^+^ and CD141^+^ cDC, while a mild downregulation of CD40 occurred on pDC (Fig. S4A). Remarkably, we observed no significant or weak associations of CD40 expression in cDCs or Mo subsets with plasma IL-6 and PCT levels with the frequency of these subsets in the blood (Fig.S4B-D). Therefore, these data highlight a differential relationship of innate activation and migration between T and NC Mo versus cDC in COVID-19 patients.

**Figure 2.**
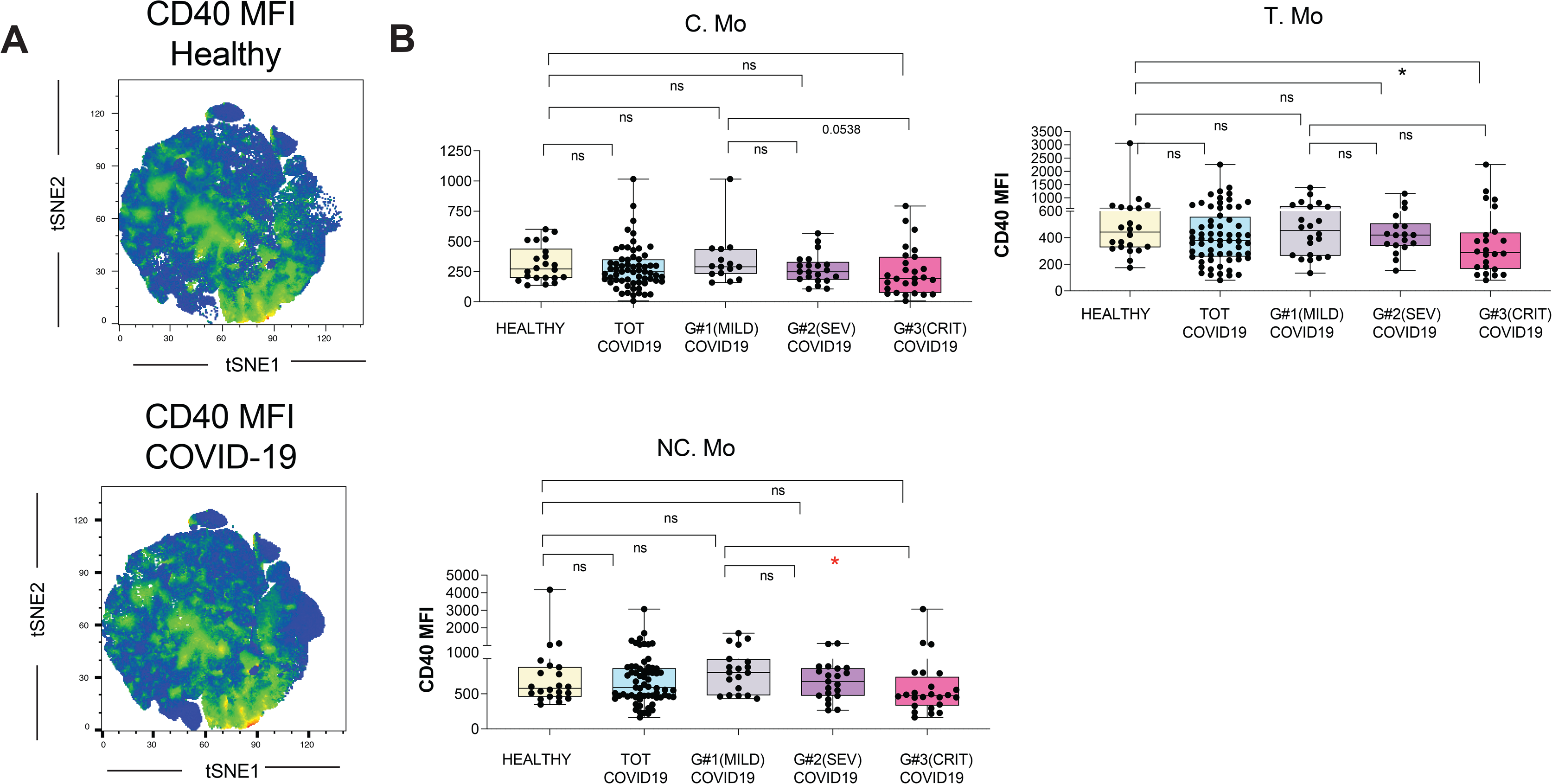
Activation profiles of myeloid cells from the blood of COVID-19 patients and association with clinical parameters. (A): tSNE plots displaying heatmaps of CD40 mean of fluorescence intensity (MFI) in different cell clusters specified in Fig. 1 from healthy control individuals (left, n = 15) and (n = 34) COVID-19 patients. (B): CD40 MFI on the indicated myeloid cell populations present in the blood of healthy individuals versus either total COVID-19 patients included in the study or patients stratified into groups according to mild (G1), severe (SEV, G2) and critical (CRIT, G3) clinical characteristics specified in Table S1. Statistical differences between patient groups were calculated using a non-parametric two tailed Mann Whitney test (black) or a Kruskal Wallis test followed by a Dunn’s post hoc-test for multiple comparisons (red). *p<0.05; **p<0.01; ***p<0.001; ****p<0.0001.

### Inflammatory transitional and non-classic Mo are enriched and activated in bronchoscopy infiltrates from COVID-19 patients with ARDS

Based on these data, we postulated that recruitment to the lung and activation of Mo and DC subsets might be different in COVID-19 patients developing severe disease and ARDS. Most studies analyzing inflammatory cell populations in the lung have used bronchoalveolar lavage (BAL) samples, since they are most representative of deep alveolar structure (24–26). However, clinical features of COVID-19 patients requiring artificial respiratory support make it difficult to collect enough BAL samples to assess significant differences in immune cell subsets. Hence, we assessed the proportions of CD45^+^ leukocytes (Fig. S5A) and their subsets (see strategy of flow cytometry gating in Fig. S5B) in dense bronchoscopy samples routinely obtained to allow patient ventilation. No significant differences in clinical parameters were observed in bronchoscopy samples with different levels of hematopoietic cell infiltrates (Fig.S5A, bottom). Remarkably, these cell distribution patterns were very similar to those observed in the few cases where a BAL sample could be obtained in parallel (Fig. S5B). Using this approach, a significant enrichment of granulocytes (which represented the vast majority of the leukocytes in those samples, FigS5B-5C), CD16^hi^ CD14^lo/-^HLA-DR^+^ cells (Fig.S5B-5C) and inflammatory T and NC Mo was observed (Fig.3A). In contrast, C Mo and CD1c^+^ DCs were also enriched but made up a very low percentage of the bronchoscopy samples. No pDCs or CD141^+^ cDCs were found in bronchoscopy infiltrates. In addition, analysis of paired blood and lung samples demonstrated that T and NC Mo, and CD1c^+^ cDCs were specifically enriched in the lung (Fig. 3B). Similar enrichment patterns were observed with granulocytes (Fig. S5D). Importantly, myeloid cells infiltrated in the lung expressed higher levels of CD40 than their circulating counterparts (Fig.3C). In the lung, we observed increasing expression of CD40 on C, T and NC Mo. The latter represented the myeloid cell subset expressing the highest levels of CD40 compared to CD1c^+^ cDC as well (Fig. 3D). In addition, levels of CD40 on NC Mo correlated with higher CRP within this group of critical COVID-19 patients (Fig. 3E). Together, these data indicate preferential enrichment inflammatory Mo in the lung and contrasting maturation profiles between infiltrated NC Mo compared to T Mo and CD1c^+^ cDCs.

**Figure 3.**
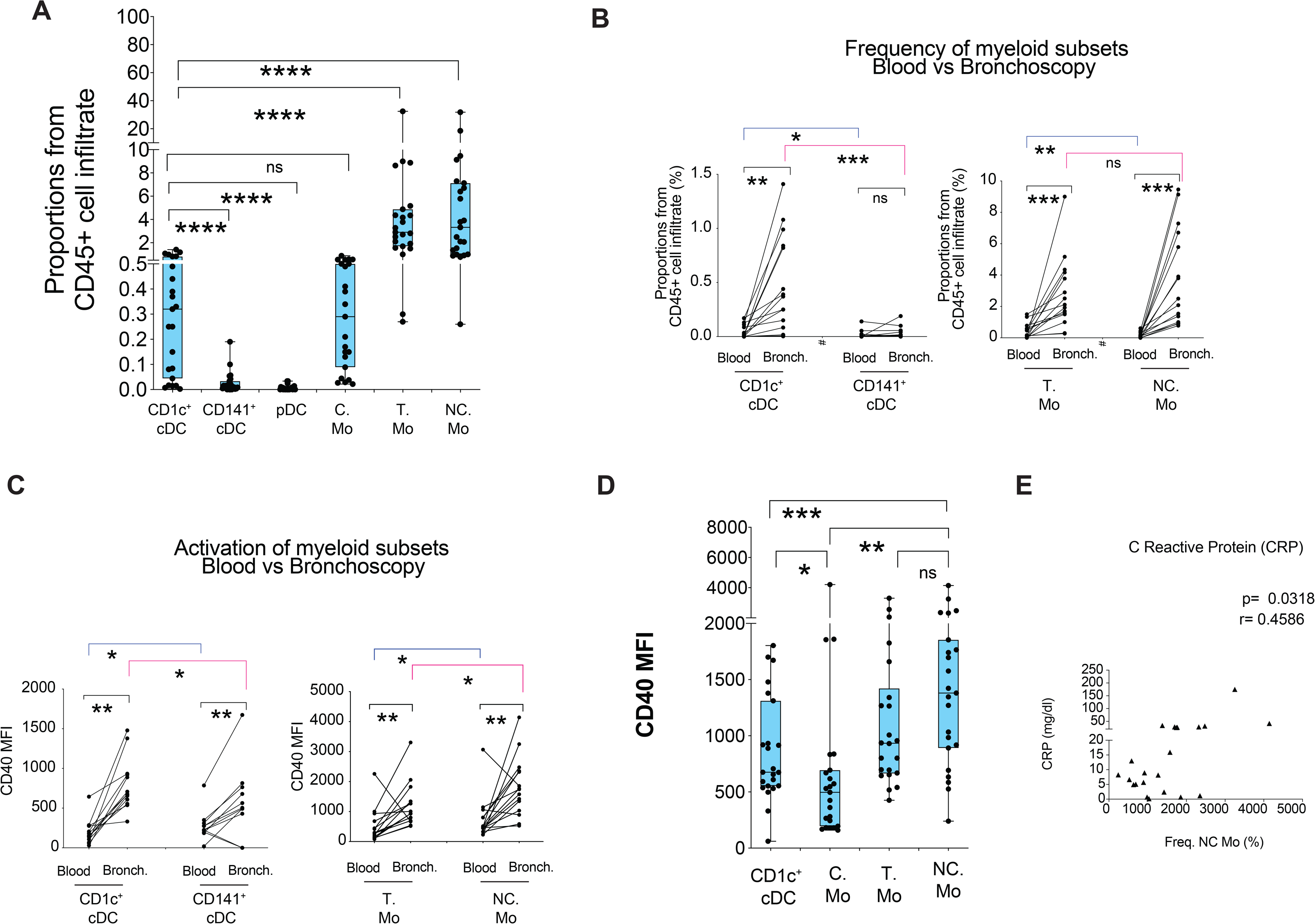
Characterization of myeloid cell subsets present in bronchoscopy infiltrates from COVID-19 patients suffering ARDS. (A): Percentages of the indicated cell populations in the hematopoietic CD45^+^ infiltrate present in bronchoscopy mucus samples from severe COVID-19 patients (n = 23) presenting ARDS and receiving IMV at ICU. Statistical differences between proportions of cell populations within the same infiltrates were calculated using a two-tailed matched pairs Wilcoxon test. (B-C) Frequencies (B) and CD40 MFI (C) of CD1c^+^ and CD141^+^ cDCs (left plot) and transitional and non-classic Mo (right plot) in paired blood and bronchoscopy samples from COVID-19 patients presenting ARDS (n = 15). Statistical significance of differences in frequencies between paired blood vs bronchoscopy samples (black) or between different cell subsets within either blood (blue) or bronchoscopy infiltrates (pink) was calculated using a two-tailed matched pairs Wilcoxon test. **p<0.01; ***p<0.001. (D): Comparison of CD40 MFI on the indicated myeloid cell populations present in the bronchoscopy infiltrates of total critical G3 COVID-19 patients. Statistical significance of differences was calculated using a two-tailed matched pairs Wilcoxon test. *p<0.05; **p<0.01. (E): Spearman correlations between C reactive protein (CRP) levels in plasma and CD40 MFI on NC Mo present in the bronchoscopy infiltrates of severe COVID-19 patients. Spearman P and R values are shown in the upper right corner of the plot.

### Effector CD8^+^ T cell populations in the lung of COVID-19 patients: Association with inflammatory Mo subsets and CD1c^+^ DC

To better understand the contribution of CD1c^+^ and inflammatory Mo to either disease progression or protective antiviral immunity in COVID-19 patients, we studied the presence of infiltrating effector T cells in bronchoscopy samples. We did not observe any significant increase of T cells or changes in the ratios of CD4^+^/CD8^+^ T cells in the lung as compared to blood (Fig. S6A). Expression of CD38 and CXCR5 in CD8^+^ T cells has been previously linked to effector function and immune exhaustion during viral infections (Fig. 4A) (27, 28). Next we analyzed expression of these activation markers in circulating CD8^+^ T cells finding a significant enrichment of activated CD38^+^ CD8^+^ T cells, more significantly those that co-expressed CXCR5 in bronchoscopies from critical COVID-19 patients requiring IMV (Fig. 4B, Fig.S6B). Of note, CD38^+^ CXCR5^+^ and CD8^+^ T cells was the only subset that was significantly altered compared to both paired blood in COVID-19 patients and to blood from healthy individuals (Fig.4B). Finally, we assessed whether frequencies or activation status of inflammatory Mo and CD1c^+^ cDCs could be associated with any of these effector CD8^+^ T cell subsets. Interestingly, we observed that higher ratios of inflammatory T and NC Mo/CD1c+ cDCs were negatively associated with proportions of CXCR5^+^ CD38^+^ CD8^+^ T cells, suggesting a detrimental role of more preferential infiltration of inflammatory Mo versus DC during disease progression (Fig. S6C). In addition, levels of CD40 levels in T Mo but no other myeloid cells in the lung were positively associated with proportions of infiltrated CD38^+^ (both CXCR5^+^ and CXCR5^−^) CD8^+^ T cells (Fig. 4C; Fig. S6D).

**Figure 4.**
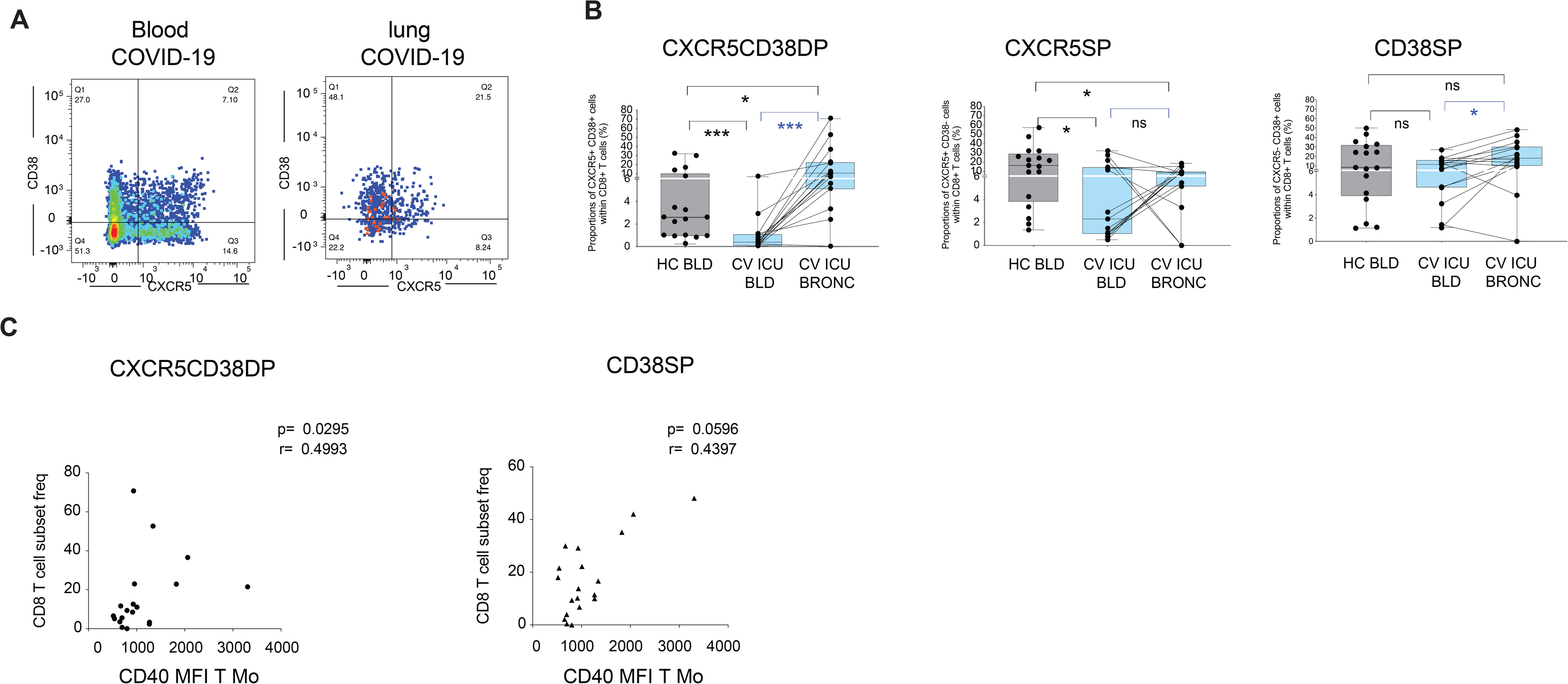
Association between effector CD8+ T cell and inflammatory myeloid cells present in bronchoscopy from COVID-19 patients with ARDS. (A): Representative flow cytometry analysis of CD38 versus CXCR5 expression on gated CD8^+^ T cells present in the blood (left) and paired bronchoscopy infiltrate (right) from a COVID-19 patients with ARDS. Numbers on quandrants represent percentage of positive cells. (B): Analysis of frequencies of CXCR5^+^ CD38^+^ (CXCFR5CD38DP; left plot), CXCR5^+^ CD38^−^ (CXCR5SP; middle plot) and CXCR5^−^CD38^+^ (CD38SP; right plot) CD8^+^ T cells present on paired blood and bronchoscopy samples from COVID-19 presenting ARDS. Frequencies of these CD8^+^ T cell subsets on the blood of healthy controls (HC) were included for reference. Statistical significance of differences in frequencies between paired blood vs bronchoscopy samples (blue) or comparison with healthy controls (black) was calculated using a two-tailed matched pairs Wilcoxon and Mann Whitney tests, respectively. **p<0.01; ***p<0.001. (C): Spearman correlations between proportions of the indicated effector CD8^+^ T cells subset and CD40 MFI on transitional (T) Mo. Spearman P and R values are shown in the upper right corner on each correlation plot.

## Discussion

In this study, associations of specific Mo and DC subsets with disease progression in COVID-19 patients are established. Our results indicate that increased proportions of transitional Mo in the blood could be used as a marker of viral control or mild clinical condition in infected patients. In contrast, dramatic decrease of T and NC Mo and CD1c^+^ DC is associated with severe clinical outcomes in COVID-19 patients. In addition, our study shows that reduction of frequencies of T and NC Mo in the blood is associated with increased IL-6, and inflammatory markers such as PCT, CRP; this reduction is also associated with the selective recruitment to the lung of these populations and CD1c^+^ cDCs during the development of ARDS. On the other hand, increased activation of NC Mo and defective maturation of T Mo might be associated with uncontrolled inflammation in the lung and the activation of effector CD8^+^ T cells. Therefore, our data highlight differential involvement of myeloid cell subsets in the pathogenesis of COVID-19 disease and immunity against SARS-Co-V2 virus. Although these data provide novel cellular insights into the pathogenesis of severe COVID-19, future studies will need to focus on mechanisms responsible for these altered cellular patterns. In this regard, recent single-cell transcriptomic analyses on inflammatory infiltrates in the lung of COVID-19 patients suggest the altered expression of genes coding for proinflammatory cytokines and/or interferons-associated and interferon-stimulated proteins that have been previously associated with the activation of antibacterial innate pathways(10).

Our data indicate that a large proportion of COVID-19 individuals from the critical G3 group developing ARDS and requiring Invasive mechanical ventilation (IMV), were superinfected with bacteria or candida at the time of sample collection. While these superinfections were mostly detected after the initiation of IMV, the majority of critical G3 COVID-19 patients exhibited high Our data indicate that a large proportion of COVID-19 individuals from the critical G3 group levels of PCT upon admission, a parameter commonly associated with bacterial infection (29) and also an indicator of inflammation and severe COVID-19 prognosis (2). Therefore, it is possible that COVID-19 patients might be more susceptible to microbial superinfection early or during ICU treatment and this could contribute to changes in inflammation, migration and homeostasis of myeloid cells from COVID-19 patients specifically undergoing severe disease progression. If this was the case, the simultaneous activation of antiviral and antibacterial innate recognition pathways could be a potential mechanism that might contribute to uncontrolled inflammation and immune exhaustion in these individuals, similarly to what has been described in HIV^+^ individuals co-infected with mycobacterium tuberculosis and developing immune reconstitution inflammatory syndrome (IRIS) (30). However, further research is required to specifically address this issue. Novel information about critical cell subsets participating in the pathogenesis of COVID-19, significant sample size and the availability of healthy non-COVID-19 controls as reference group are strengths of our research. However, a number of limitations need to be discussed. This is a cross-sectional study design comparing severe COVID-19 patients, with two sets of controls: on the one hand mild/severe COVID-19 patients; and on the other hand non-COVID-19 healthy controls. Because of this cross-sectional nature any assessment of progression will require a confirmation in prospective follow-up studies. In addition, our study provides data on bronchoscopy infiltrates rather than bronchoalveolar lavage given the fragile status of COVID-19 patients with ARDS. Although bronchoscopies might not necessarily reflect the characteristics of infiltrates present in terminal bronchioles and alveoli, our data from two patients indicate that the composition of myeloid cells in the infiltrates obtained by these methods are comparable. On the other hand, our study did not directly address the protective versus detrimental function of DC during COVID-19 infection. In this sense, CD141^+^ cDCs are important for the priming of antiviral exhaustion in these individuals, similarly to what has been described in HIV^+^ individuals co-CD8^+^ T cell responses (14, 31) and are dramatically depleted from the blood in all COVID-19 patients regardless of their clinical status, but they are almost completely undetectable in bronchoscopy infiltrates from individuals developing severe ARDS. These data might support a defect on the priming, the maintenance or exhaustion of virus-specific CD8^+^ T cells in the lungs; however these issues must be investigated in depth. In addition, CD1c^+^ cDCs are known to support CD4^+^ T cell responses and stimulate follicular helper T cells required for effective humoral antiviral adaptive immunity (13). While it has been proposed that protection against SARS-CoV-2 might also be at least partially mediated by specific antibodies (32), the role of CD1c^+^ DCs inducing Tfh responses in these patients has not been addressed in our study and requires further research.

To conclude, our study unveils immune cellular networks associated with control or progression of COVID-19 disease, which might be useful for early diagnosis, and the design of new more targeted and individualized treatments. Eventually, further research on these mechanisms may lead to the development of new therapeutic or preventive strategies.

## Material and Methods

### Study patient cohort and samples

A total of 64 patients diagnosed with COVID-19 after testing positive for SARS-CoV-2 RNA by qPCR were included in the study. As specified in Supplemental Table 1, 95% of patients initiated different treatments upon hospital admission. Our cohort of COVID-19 patients was stratified into 3 groups of mild (G1), severe (G2) and critical (G3) prognosis based on respiratory frequency (RF), blood oxygen saturation (StO2), partial pressure of arterial oxygen to fraction of inspired oxygen ratio (PaFiO2) and respiratory failure values as well as by following recently described criteria (21).

Blood samples were obtained for all participants and directly used for analysis without prior ficoll gradient centrifugation, to avoid the depletion of polymorphonuclear cells.

Of note, critical G3 patients from which blood (n = 24) and/or bronchoscopy samples (n = 23) were collected displayed 85.71% and 100% of microbial superinfection at the time of sample collection, respectively. The majority of superinfections in these patients were observed 6 days after ICU admission and IMV support (89% from blood and 100% from bronchoscopy samples) and were caused by either only bacteria (68.75% from blood and 65% from bronchoscopy samples) or only fungi (25% from blood and 30% from bronchoscopy samples) or both types of parthogens (6.25% from blood and 5% from bronchoscopy samples) (Table S2).

Flexible bronchoscopy procedures were conducted in all ICU patients under sedation and muscle relaxation. This technique enables the visualization of the lumen and mucosa of the trachea, and also of the proximal and distal airways. Flexible bronchoscopy is indicated to evaluate pneumonia, to asses infiltrates of unclear etiology, to aspirate secretions, and to prevent development of atelectasis and pneumonia associated with invasive ventilation. Furthermore, in patients suspected of having pneumonia or infections, microbiological specimen can be collected with bronchial washings. Bronchial washings consist of aspirating bronchial secretions directly or after instilling physiological serum. We did not employ local anesthetic to avoid sample alterations. Bronchoalveolar lavage can also be performed in pneumonia patients. Nevertheless, this technique entails a high risk for patients with severe respiratory failure. This is true specially for patients with high oxygen concentration requirements such as COVID pneumonia patients. Therefore, we performed bronchoalveolar lavage solely in two cases (33). Finally, the hematopoietic lung infiltrates were obtained from bronchoscopy samples diluted in 0.9% sodium chloride at a 1: 5 ratio and after three sequential centrifugations at 1500 rpm during 10 minutes.

### Flow cytometry reagents and data analysis

For phenotypical studies of cell populations present in peripheral whole blood, cells were incubated with a panel of different combinations of the following monoclonal antibodies: anti-human CD3-Pacific blue and PerCP-Cy5.5, CD19, CD56 PerCP-Cy5.5-, CD8-APC-Cy7, CXCR5-PE, CD14-PE, CD16-Pacific Blue, CD40-FITC, HLA-DR-APC-Cy7 and –APC, CD11c-Pacific Blue, CD1c-PE-Cy7, CD141-APC (Biolegend) and CD38-FITC, CD123-APC, CD20-PerCP_Cy5.5 and HLA-DR-APC (BD Becton Dickinson) for 30 min. Subsequently, whole blood cells were treated with BD FACS Lysing Solution for 15 min, centrifuged and finally resuspended in PBS 1X (LONZA) and analyzed using a BD FACSCanto II flow cytometer (BD Becton Dickinson). The gating strategy for myeloid cells is shown in Fig. S2: CD1c^+^ and CD141^+^ cDC subsets were identified from as Linage negative (CD3^−^ CD19^−^ CD20^−^ CD56^−^) negative, CD14- HLA^−^DR^+^ differing on exclusive expression of CD1c and CD141 (panel I). Plasmacytoid DCs(pDC) were defined as Lin^−^HLA-DR^+^ CD11c^−^ CD123^hi^ cells (Panel II). CD14^hi^ CD16^−^ Classical(C), CD14^int^ CD16^+^ transitional (T) and CD14^low^ CD16^hi^ non-classical (NC) monocytes were defined on the basis of CD14 and CD16 expression levels. Granulocytes were identified as large CD14^lo/-^CD16^hi^ HLADR^−^ cells. Maturation status of myeloid cells was defined based on CD40 mean fluorescence intensity (MFI). In the case of bronchoscopy samples from critical COVID-19 patients, we added anti-CD45-Pacific Orange mAbs (BioLegend) to the previously described antibody cocktails and performed an additional panel to analyze frequencies and distribution of CD3^+^ CD8^+^ and CD4^+^ T cell subsets defined by expression of CD38 and CXCR5 or HLA-DR, respectively. Individual and multiparametric analysis of flow cytometry data was performed using flowJo software (Tree Star) and tSNE.

### Statistical Analysis

Quantitative variables were represented as median and interquartile range. Statistical differences between different cell populations or between patient cohorts between was calculated using a nonparametric two-tailed Mann Whitney test, or using a Kruskal wallis test followed by a multiple comparisons, as appropriate. Significant differences of paired analyses were calculated using a two-tailed Wilcoxon matched pairs test and a Bonferroni multiple comparison correction when possible. Association between cellular and clinical parameters was calculated using nonparametric Spearman correlations. Statistical significance was set a p<0.05. Statistical analyses were performed using the GrapPad Prism 8 Sofware.

### Ethics

This study was approved by the Research Ethics Committee from Hospital Universitario de La Princesa in the context of REINMUN-COVID and EDEPIMIC projects and it was carried out following the ethical principles established in the Declaration of Helsinki. All included patients (or their representatives) were informed about the study and gave written informed consent.

## Data Availability

Nothing to declare

## Author contributions

E.M.G., J.A., J.B.S., F.S.M, C.M.C, I.S.C and P.L developed the research idea and study concept, designed the study and wrote the manuscript;

E.M.G., J.A. C.M.C, A.A and JBS supervised the study;

I.S.C. and P.L. designed and conducted most experiments and equally contributed to the study; J.A., J.B.S and P.L. provided Peripheral blood and bronchoscopy samples from study patient cohorts

All other authors participated in patient samples processing and the creation of a clinical data database used for the study.

## Supplementary Materials

**Fig. S1.**
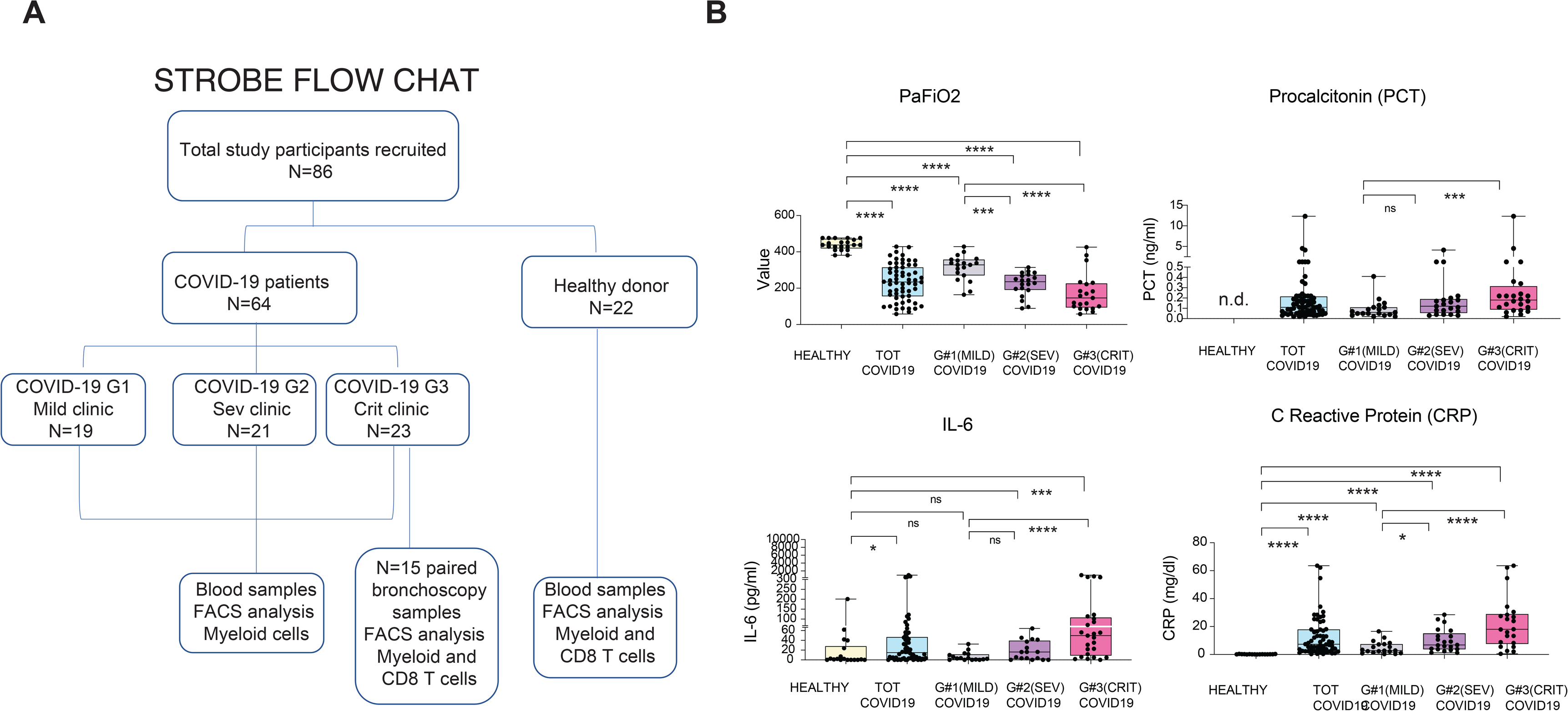
Selected clinical parameters differentially detected in subgroups of COVID-19 patients with different clinical severity. (A): STROBE Flow chart of the study showing patient recruitment and stratification according to clinical severity (B): Clinical values of PaFiO2, IL-6, procalcitonin, and C Reactive Protein detected in the blood of total number of COVID-19 patients or patientsstratified into mild (G1), severe (G2) and critical (G3) clinical status defined according to the criteria detailed in Table S1. Values present in healthy controls (HC) were included when available. Statistical significance of differences in clinical values across the study groups were calculated using a Mann Whitney or Kruskal wallis and Dunn’s post-hoc tests. *p<0.05; **p<0.01; ***p<0.001; ****p<0.0001.

**Fig. S2.**
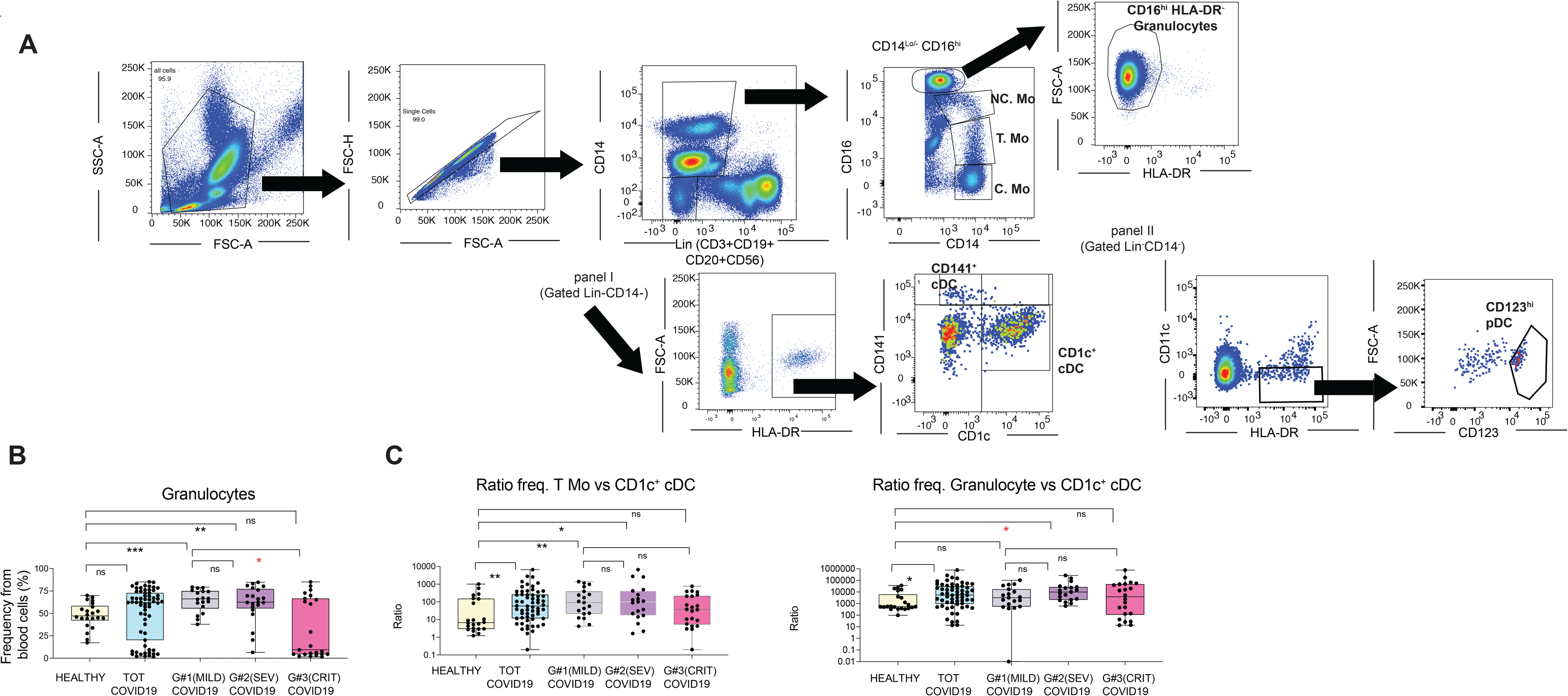
Flow cytometry characterization of myeloid cell subsets from COVID-19 patients. (A): Flow cytometry gating strategy showing Monocyte (Mo), conventional (cDC) and plasmacytoid DC (pDC) and granulocyte characterization in the blood from a representative patient of our study cohorts. CD14^+^ Lineage (CD3^−^ CD19^−^ CD20^−^ CD56^−^) negative Mo subsets were defined by CD14 and CD16 expression levels. Granulocytes were identified as large CD16^hi^CD14^lo/-^HLADR^−^ cells. cDC subsets were identified as CD14^−^ Lin^−^ HLA-DR^+^ big cells differing on CD1c and CD141 expression. Finally, pDCs were defined as CD14^−^Lin^−^CD11c^−^ HLADR^+^ CD123^hi^ lymphocytes. (B-C): Proportions of granulocytes (B) and ratios of frequencies between Transitional Mo/CD1c^+^ cDC and Granulocyte/ CD1c^+^ cDCs (C) present in the blood of healthy individuals compared with either total COVID-19 patients included in the study or patients stratified into mild (G1), severe (SEV, G2) and critical (CRIT, G3). Statistical differences between patient groups were calculated using a non-parametric two tailed Mann Witney test (black) or a Kruskal Wallis test followed by a Dunn’s post hoc-test for multiple comparisons (red). *p<0.05; **p<0.01; ***p<0.001.

**Fig. S3.**
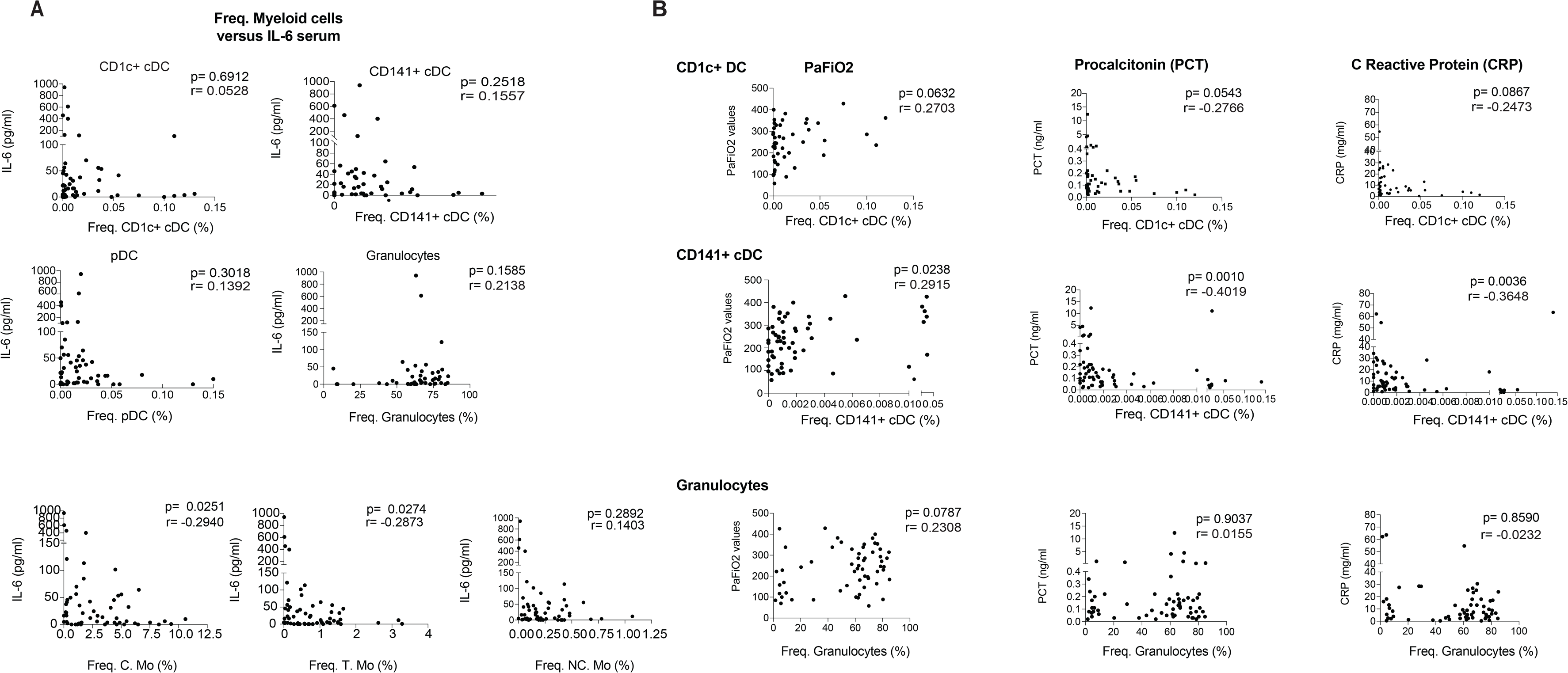
Correlation of frequencies of circulating myeloid cell subsets with IL-6 plasma levels. Spearman correlations between IL-6 plasma levels (A) or inflammatory clinical values (B) and percentages of the indicated myeloid cell subsets in the blood of COVID-19 patients. Spearman P and R values are shown in the upper right corner of the plot.

**Fig. S4.**
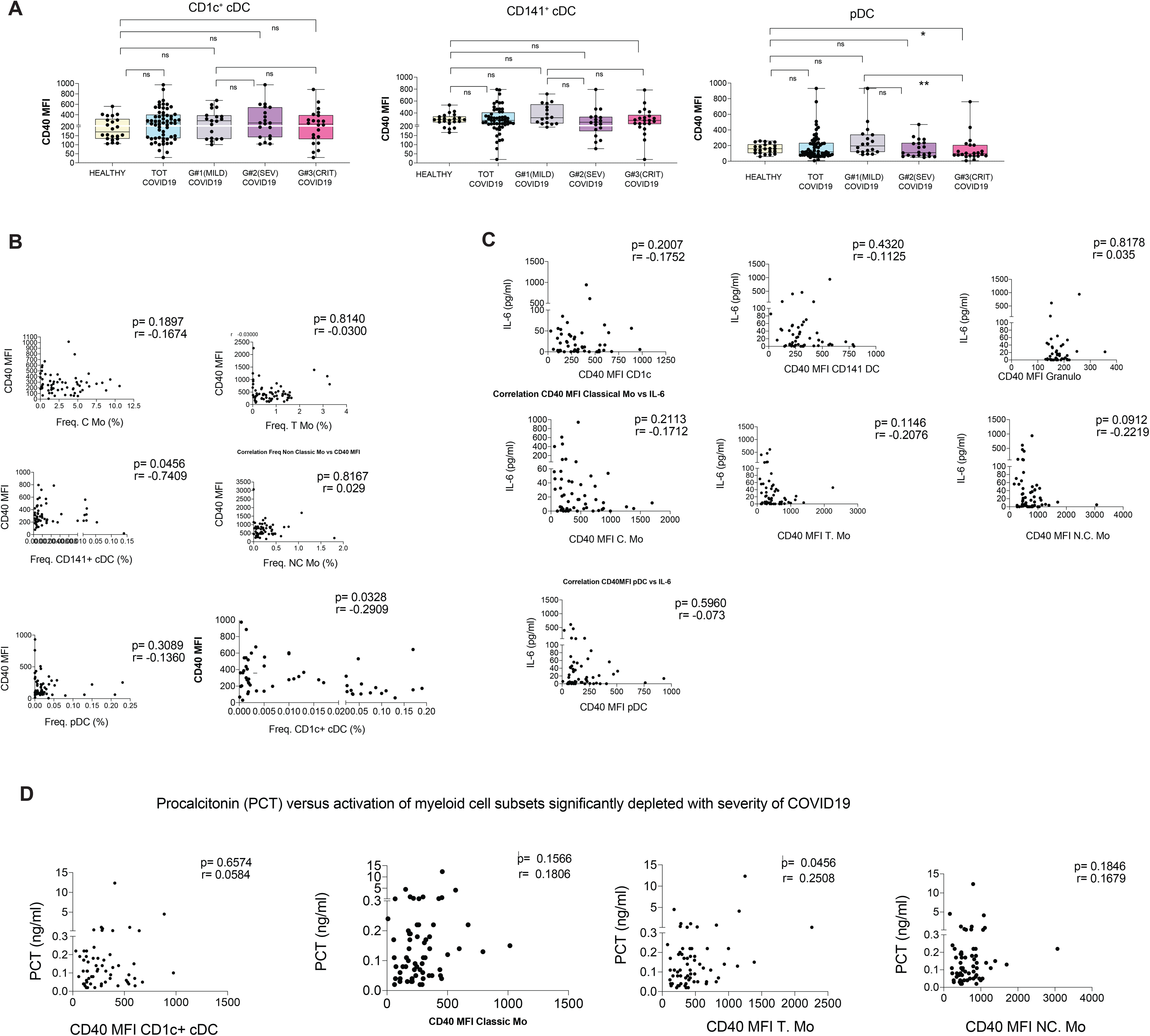
Correlation between activation status of myeloid cell subsets and frequencies in the blood and IL-6 plasma levels. (A): CD40 Mean of fluorescence intensity (MFI) on the indicated myeloid cell populations present in the blood of healthy individuals versus either total COVID-19 patients included in the study or patients stratified in mild (G1), severe (SEV, G2) and critical (CRIT, G3) clinical characteristics specified in Table S1. (B-C): Spearman correlation between CD40 MFI on the indicated myeloid cell subsets and their frequency in blood (B) or IL-6 plasma levels (C). Spearman P and R values are shown in the upper right corner of the plot. Statistical significance of each cell subset between each patient subgroup was tested using a two tailed Mann Whitney test. (D): Spearman correlations between procalcitonin (PCT) levels in plasma and CD40 mean fluorescence intensity (MFI) on CD1c+ cDCs,classical (C), transitional (T) and non-classical (NC) Mo. Spearman P and R values are shown in the upper right corner of the plot.

**Fig. S5.**
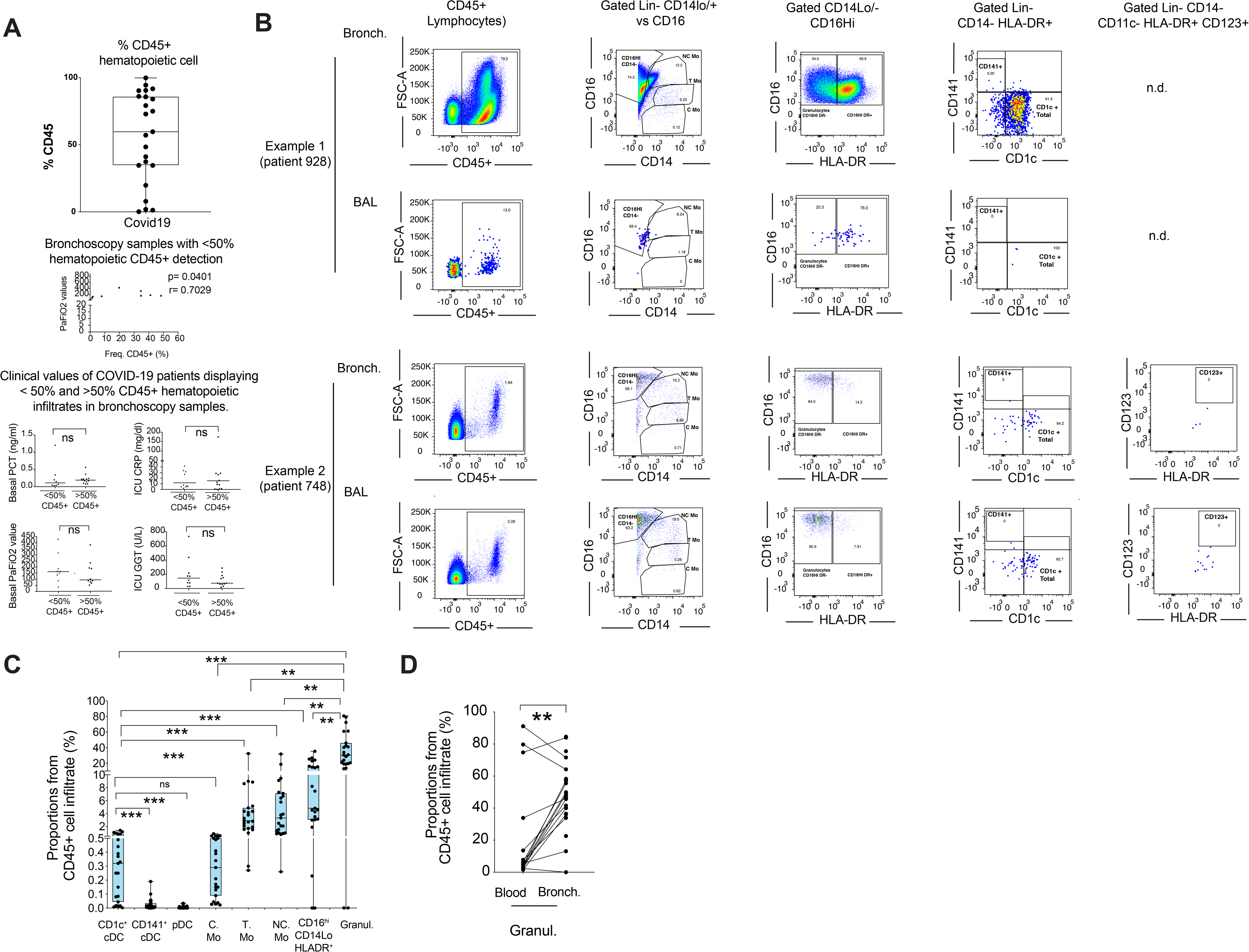
Characterization of myeloid cells present in bronchoscopy infiltrates from COVID-19 patients with ARDS. (A): Proportions of CD45^+^ hematopoietic cells present in bronchoscopy samples obtained from COVID-19 patients (n = 23) with severe ARDS and requiring IMV at hospital’s ICU. Associations with clinical parameters in samples with frequencies of CD45+ hematopoietic cell superior or inferior to 50% are shown below. (B): Flow cytometry analyses of myeloid cell subsets hematopoietic infiltrates from bronchoscopy compared with bronchoalveolar lavage (BAL) from the same COVID-19 severe patient. Two representative examples are shown. (C): Percentages of the indicated cell populations in the hematopoietic CD45^+^ infiltrate present in bronchoscopy mucus samples from severe COVID-19 patients (n = 23) presenting ARDS and receiving IMV at ICU. Statistical differences between proportions of cell populations within the same infiltrates were calculated using a two-tailed matched pairs Wilcoxon test. (D): Frequencies of CD14lo/- CD16hi HLA-DR-granulocytes on the blood and paired bronchoscopy samples from critical COVID-19 patients. Statistical differences were calculated using a two-tailed matched pairs Wilcoxon test. **, p<0.01; ***;p<0.001.

**Fig. S6.**
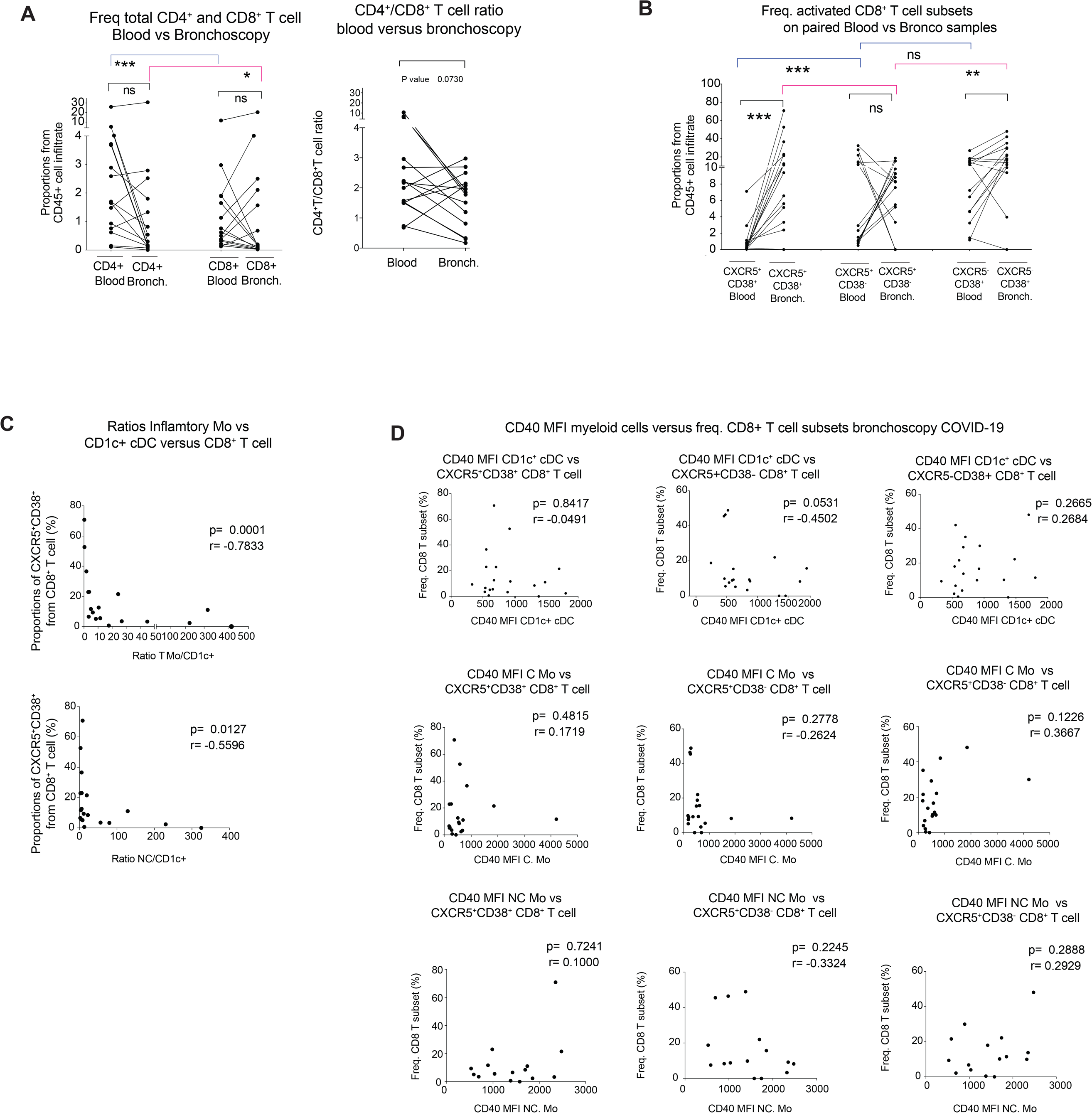
Characterization of effector CD8+ T cells present in bronchoscopy infiltrates from COVID-19 patients with ARDS. (A-B): Percentages of total CD4^+^ and CD8^+^ T cells and CD4^+^T/CD8^+^T ratios (A) or CXCR5^+^CD38^+^ (DP), CXCR5^+^ CD38^−^ (CXCR5SP) and CXCR5^−^CD38^+^ (CD38SP) (B) from CD8^+^ T cells in the blood and paired bronchoscopy samples from COVID-19 patients with ARSD (n = 15). Statistical significance was calculated using a two tailed matched pairs Wilcoxon test for differences of the same populations between different tissue localizations (black) or between different cell populations included within the blood (blue) or within the bronchoscopy (pink) samples. (C-D): Spearman correlations between proportions of CXCR5^+^CD38^+^, CXCR5^+^ CD38^−^ and CXCR5^−^ CD38^+^ subpopulations of CD8^+^ T cells from bronchoscopy samples from COVID-19 patients and ratios of frequencies between Transitional(T) or non-classic (NC) Mo versus CD1c^+^ cDCs (C) and CD40 MFI of the indicated myeloid populations in bronchoscopy samples (D). Spearman P and R values are shown in the upper right corner of the plot.

**Table S1.** Demographic and clinical information from study patient cohorts.

|  | Healthy | All COVID-19 Blood | G1 Mild COVID-19 Blood | G2 Severe COVID-19 Blood | G3 Critical COVID-19 Blood | G3 Critical COVID-19 Bronch. |
| --- | --- | --- | --- | --- | --- | --- |
| Number of patients (n) | 22 | 64 | 19 | 21 | 24 | 23 |
| Median time since admission until sample collection (*days, Min-Max*) | n.d. | 3 (0-25) | 1 (0-5) | 2 (0-23) | 11 (1-25) | 14 (3-25) |
| Duration of symptoms at sample collection | n.d. | 8.5 (0-30) | 9 (2-20) | 8 (0-15) | 7 (1-30) | 19 (1-30) |
| Median Age (*years, Min-Max*) | 52 (23-82) | 61 (22-89) | 61 (22-86) | 58 (27-89) | 61 (28-75) | 61 (28-75) |
| Sex (% Male) | 22.73% | 57.8% | 47.37% | 57.14% | 66.67% | 61% |
| Clinical parameters (*median, min-max*) |  |  |  |  |  |  |
| Respiratory rate (rpm) | 14 (12-20) | 24 (12-49) | 20 (12-26) | 24 (14-40) | 34 (20-49) | 32 (15-49) |
| PaFiO2 | 438.09 (380.95-476.2) | 233 (58-428.6) | 328.09 (163.89-428.6) | 228.57 (89-314.28) | 146.25 (58-426) | 114.8 (70-426) |
| Comorbidities (n,%) | 2 missing |  |  |  |  |  |
| Tobacco | 4 (20%) | 11 (17.46%) | 0 (0%) | 7 (35%) | 4 (16.67%) | 6 (26.09%) |
| Hypertension | 3 (15%) | 18 (28.13%) | 3 (15.79%) | 6 (28.57%) | 9 (37.5%) | 10 (43.48%) |
| Diabetes Mellitus | 1 (5%) | 12 (18.75%) | 1 (5.26%) | 3 (14.29%) | 8 (33.33%) | 6 (26.09) |
| Dyslipidemia | 3 (15%) | 22 (34.48%) | 4 (21.05%) | 9 (42.86%) | 9 (37.5%) | 10 (43.48) |
| History of cardiovascular disease | 0 | 9 (14.06%) | 1 (5.26%) | 3 (14.29%) | 5 (20.83%) | 5 (21.74%) |
| History of lung disease | 3 (15%) | 13 (20.31%) | 4 (21.05%) | 6 (28.57%) | 3 (12.5%) | 7 (30.43%) |
| Laboratory values (*median, min-max*) |  |  |  |  |  |  |
| Serum IL-6 (pg/ml) | 1.5 (0-201) | 14.86 (0-941.26) | 3.69 (0-53.54) | 16.41 (0-64.37) | 49.8 (0-941.26) | 47.4 (2.6-458.5) |
| PCT (ng/ml) | n.d. | 0.11 (0.02-12.34) | 0.06 (0.02-0.41) | 0.12 (0.03-4.14) | 0.18 (0.02-12.34) | 0.18 (0.02-1.2) |
| CRP (mg/dl) | 0.11 (0-0.42) | 7.31 (0.32-63.6) | 2.85 (0.32-16.57) | 6.8 (1.19-28.5) | 18.02 (0.47-63.6) | 18 (0.47-63.6) |
| Glucose (mg/dl) | 89 (69-142) | 122 (73-327) | 105 (77-174) | 113 (73-190) | 150 (79-327) | 125 (95-327) |
| GGT (U/L) | 13.5 (7-40) | 62 (12-1743) | 52 (15-247) | 62 (14-1743) | 63 (12-609) | 60 (21-609) |
| Lymphocyte Count | 2275 (1150-4710) | 1030 (160-7200) | 1230 (240-2310) | 1060 (330-2930) | 670 (160-7200) | 980 (210-7200) |
| D-dimer (µg/ml) | n.d. | 0.8 (0.22-42.1) | 0.63 (0.24-5.66) | 0.79 (0.31-6.68) | 2.13 (0.22-42) | 0.9 (42-0.3) |
| LDH (U/L) | 154 | 300 | 236 | 289 | 432 | 448 |

**Supplemental Table 1. Demographic and clinical information from study patient cohorts.**
|  | (134-179) | (119-1319) | (119-236) | (184-474) | (244-1319) | (252-1319) |
| --- | --- | --- | --- | --- | --- | --- |
| Ferritin (ng/ml) | 72<br>(10-325) | 1111<br>(124-8266) | 753<br>(124-4234) | 1097<br>(232-3474) | 1555<br>(442-8266) | 1000<br>(7468-274) |
| **Intercurrent events** |  |  |  |  |  |  |
| Superinfection *n* (%) | 0 | 20 (32.79%)<br>3 missing | 0 | 2 (9.52%) | 18 (85.71%)<br>3 missing | 20 (100%)<br>3 missing |
| Treatment upon admission to<br>extraction blood/bronco *n* (%) | 0 | 61 (95.31%) | 16 (84.21%) | 21 (100%) | 22 (91.67%) | 23 (100%) |
| Hidroxychloroquine |  | 61 (95.31%) | 16 (84.21%) | 21 (100%) | 22 (91.67%) | 23 (100%) |
| Lopinavir/Ritonavir |  | 50 (78.13%) | 13 (68.42%) | 17 (80.95%) | 18 (75%) | 19 (82.6%) |
| Azithromycin |  | 55 (85.94%) | 16 (84.21%) | 17 (80.85%) | 22 (91.66%) | 21 (91%) |
| Ceftriaxone |  | 31 (48.44%) | 2 (10.53%) | 10 (47.62%) | 19 (79.16%) | 15 (65%) |
| Other antibiotics |  | 18 (28.57%) | 0 | 4 (19.05%) | 14 (58.33%) | 20 (87%) |
| Glucocorticoids |  | 35 (54.69%) | 8 (42.11%) | 8 (38.1%) | 19 (79.17%) | 21 (91%) |
| Tozilizumab |  | 19 (29.69%) | 0 | 3 (14.29%) | 16 (66.67%) | 18 (78%) |

**Table S2.** Detection of microbial superinfection in severe and critical COVID-19 patients.

|  | G2 Severe<br>COVID-19 | G3 Critical<br>COVID-19<br>Blood | G3 Critical<br>COVID-19<br>Bronch. |
| --- | --- | --- | --- |
| **Number of patients n, (%)** | **2, 9.52%** | **18, 85.71%** | **23, 100%** |
| Time upon admission until ICU<br>(days, median min-max) | n.d. | 0 (0-12) | 3 (0-12) |
| Duration of symptoms until extraction<br>blood/broncho<br>(days, median min-max) | 2.5 (0-5) | 10 (2-30) | 11 (5-31) |
| Time upon admission ICU to culture<br>extraction (days, median min-max) | 7 (6-8) | 6 (0-20) | 6 (0-16) |
| PCT (ng/ml) (median min-max) | 0.12<br>(0.11-0.12) | 0.22<br>(0.04-12.34) | 0.2<br>(0.03-1.2) |
| ICU n, (%) | 0 | 17 (94.44%) | 23 (100%) |
| **IMV n, (%)** | **0** | **16 (89%)** | **23 (100%)** |
| **NO IMV n, (%)** | **2** | **2 (11%)** | **0** |
| **Type of superinfection in IMV n, (%)** |  |  | **3 missing** |
| Bacterial | n.d. | 11 (68.75%) | 13 (65%) |
| Fungal | n.d. | 4 (25%) | 6 (30%) |
| Both | n.d. | 1 (6.25%) | 1 (5%) |
| **Type of superinfection in NO IMV**<br>**n, (%)** |  |  |  |
| Bacterial | 2 (100%) | 2 (100%) | n.d. |
| Fungal | 0 | 0 | n.d. |
| Both | 0 | 0 | n.d. |
| **Organ of superinfection n, (%)** |  |  | **3 missing** |
| Lung | 1 (50%) | 8 (50%) | 11 (55%) |
| Periferical blood | 1 (50%) | 4 (25%) | 4 (20%) |
| Urinary | 0 | 2 (12.5%) | 2 (10%) |
| Two organs | 0 | 2 (12.5%) | 3 (15%) |

